# Higher levels of serum IL-1β and TNF-α are associated with an increased probability of major depressive disorder

**DOI:** 10.1101/2020.10.04.20206508

**Authors:** Rajesh Das, Md. Prova Zaman Emon, Mohammad Shahriar, Zabun Nahar, Sardar Mohammad Ashraful Islam, Mohiuddin Ahmed Bhuiyan, Sheikh Nazrul Islam, Md. Rabiul Islam

## Abstract

Major depressive disorder (MDD) is a serious psychiatric disorder but there are no reliable risk assessment tools for this condition. The actual reason for affecting depression is still controversial. It is assumed that the dysregulated cytokines are produced due to the hyperactivation of the immune system in depression. We aimed to evaluate the possible alteration and the role of serum interleukin-1β (IL-1β) and tumor necrosis factor-α (TNF-α) in MDD patients. The diagnostic and statistical manual of mental disorders, 5^th^ edition was used to diagnose patients and evaluation of healthy controls (HCs). The severity of depression was measured by the Hamilton depression rating scale (Ham-D). Serum IL-1β and TNF-α levels were quantified by enzyme-linked immunosorbent assay kits. Increased levels of serum IL-1β and TNF-α were observed in MDD patients compared to HCs. These higher levels of peripheral markers were positively correlated with the severity of depression. Moreover, females with higher Ham-D scores showed greater serum IL-1β and TNF-α levels compared to males. Good predictive values were detected for both serum IL-1β and TNF-α levels by receiver operating characteristic analysis. Therefore, the elevated levels of serum IL-1β and TNF-α might be used as risk assessment indicators for MDD.

## 1. Background

Major depressive disorder (MDD) is a serious public health concern due to the high prevalence, longer duration, or recurrence with the ability to impair an individual’s personal and work life (Vilagut et al., 2016). The distinctive symptoms of MDD are empty feeling, the hopeless, sad, anxious, helpless, worthless, ashamed, restless, guilty, or suicidal tendency (Kim et al., 2015). There are several hypotheses postulated for the understanding the pathogenesis of MDD such as genetic, biological, social, environmental, and psychological (Belujon and Grace 2014; Emon et al., 2020; Grassi-Oliveira et al., 2011; Hasin et al., 2005; Kessler et al., 2005). Biological cause of depression has different hypotheses like serotonin hypothesis; monoamine hypothesis where lack of monoamines is observed (Belujon and Grace, 2014, Cabib and Puglisi-Allegra, 2012; Thompson et al., 2015). The Hypothalamic pituitary adrenal (HPA) axis plays a major role in the development of depression. Increased level of inflammatory cytokine alters the metabolism of dopamine, serotonin, and noradrenaline results in the activations of the HPA axis which ultimately cause depression (Pariante and Lightman, 2008). The accuracy of the diagnosis of depression by the diagnostic and statistical manual of mental disorders (DSM) is a major concern. Also, the question format undergoes tremendous revision and the implementation of the new DSM is a great challenge for the clinicians (Patten et al., 2015). Thus, any precise diagnostic or early risk assessment markers are required for major depression however these sorts of tools are still absent in this field.

Interleukin-1β (IL-1β) and tumor necrosis factor-α (TNF-α) are pleomorphic cytokines. Therefore, serum levels of these factors may vary from person to person due to pathological states (Johnston et al., 2002; Sarkar et al., 2018). A conflict is raised about the concentration of IL-1β and TNF-α in MDD patients whether the concentration is increased or decreased. In recent years, tremendous research is going on for the establishment of accurate biological risk assessment markers for depression. Many researchers reported the altered levels of certain biological markers both proinflammatory cytokines and chemokines may be responsible for MDD (Cassano et al., 2017; Lopresti et al., 2013; Schmidt et al., 2011). Some study observed the elevated serum levels of IL-1β, TNF-α, cortisol, CRP, IL-6, IL-10, IL-15, IL-18, and MDA in MDD patients while others showed no alterations (Fan et al., 2017; Islam et al., 2018; Nishuty et al., 2019; Ogłodek et al., 2017; Rivera et al., 2014; Uguz et al., 2015; Yang et al., 2007). Some studies demonstrated positive correlations between these markers and severity of depression (Dahl et al., 2014; Dowlati et al., 2010; Farooq et al., 2017; Grassi-Oliveira et al., 2011; Haapakoski et al., 2015) whereas others reported a negative association between them (Einvik et al., 2012; Rief et al., 2010). Moreover, several studies observed no such relationships for serum IL-1β, CRP, and TNF-α levels (Brambilla and Maggioni, 1998; Marques-Deak et al. 2007; Nishuty et al., 2019). Taken all these together, the present study intended to assess the role of serum IL-1β and TNF-α in major depression.

## 2. Methods

### 2.1. Study population

This prospective case-control study enrolled 87 MDD patients and 87 healthy controls (HCs). The patients were recruited from Bangabandhu Sheikh Mujib Medical University (BSMMU), Dhaka, Bangladesh, and HCs from different parts of Dhaka city matched by socio-demographic profiles e.g. age, sex, body mass index (BMI), etc. We included drug-free newly diagnosed MDD patients for this study who were suffering from MDD symptoms for at least two weeks or longer. MDD patients were taken for this study after skillful diagnosis by a qualified psychiatrist according to DSM, 5^th^ edition (DSM-V). The severity of depression was measured by the Hamilton depression rating (Ham-D) scale and all the patients had a Ham-D score greater than 7. A pre-designed structured questionnaire was applied to record the socio-demographic and biographical profiles of the study population. Exclusion criteria include mental retardation, comorbid psychiatric illness or other serious neuropsychiatric disorders, abnormal BMI, immune disorders, and the presence of infectious diseases, alcohol and narcotic drug dependency and were ensured to be free from taking any other medication that could interfere with the serum levels of the parameters.

The ethical review committee of the department of psychiatry, BSMMU, approved this study protocol. All investigations were conducted as per the guideline stated in the Declaration of Helsinki. Before participation, a detailed brief about the aim of this study was given to each participant and written consent was taken from them.

### 2.2. Blood collection and serum separation

Five-milliliter blood sample was collected from the cephalic vein of all study participants at morning between 8 to 9 AM after overnight fasting. The collected samples were allowed to clot for one hour at room temperature for the proper separation of blood cells and serum. These clotted blood samples were then centrifuged at 1000 g for 15 min to extract the serum samples. The separated serum samples were carefully stored in -80° C until further analysis.

### 2.3. Quantification of serum IL-1β and TNF-α levels

Serum levels of IL-1β and TNF-α in the study population were analyzed by commercially available enzyme-linked immunosorbent assay (ELISA) kits (Abcam, USA). The sensitivity or minimum detectable dose for both the cytokines was less than 2 pg/mL and there was no cross-reactivity with the other cytokines present in the serum sample. The assay procedure was performed as per the instruction of the manufacturer of kits.

### 2.4. Statistical analysis

To determine the potential difference between the groups, the Wilcoxon-Mann-Whitney test was applied for categorical variables and Fisher’s exact test for non-categorical variables. Data were presented as mean ± standard deviation (SD). Mann Whitney U test and Spearman’s rank correlation test were applied to find significant changes as the data distribution was nonparametric. Variations among data were visually demonstrated by error bars and scatter plot graphs. The receiver operating characteristic (ROC) curve was drawn for the determination of the cut-off point and diagnostic performance of analyzed cytokines. Results were considered significant with *p* values less than 0.05. All the statistical analyses were performed by the statistical package for social science (SPSS) version 25.0 (IBM Corporation, Armonk, New York, USA).

## 3. Results

Significant variations in terms of age, gender, and BMI were not observed between the groups. Besides, we found no differences in education, occupation, economic status, and smoking habit between MDD patients and HCs (Table 1). According to the present study findings, most of the MDD patients were female (63%), jobless (52%), tobacco user (69%), and belonging to the medium economic status. The percentage of guilty feeling and a suicidal tendency among MDD patients were 61% and 59%, respectively.

**Table 1.** Socio-demographic characteristics and clinical information of the study population.

| Variables | MDD patients (n = 87)<br>Mean ± SD | Healthy controls<br>(n = 87)<br>Mean ± SD | *P value* |
| --- | --- | --- | --- |
| Age | 31.33 ± 9.36 | 30.46 ± 8.41 | 0.674 |
| Gender |  |  | 0.356 |
| Male | 32 (37%) | 37 (43%) |  |
| Female | 55 (63%) | 50 (57%) |  |
| BMI (kg/m<sup>2</sup>) | 23.22 ± 4.26 | 23.28 ± 3.65 | 0.945 |
| Years of education | 11.83 ± 3.38 | 12.75 ± 4.11 | 0.334 |
| Occupation |  |  | 0.147 |
| Service | 10 (11%) | 10 (11%) |  |
| Business | 14 (16%) | 11 (13%) |  |
| Student | 13 (15%) | 13 (15%) |  |
| Jobless | 45 (52%) | 38 (44%) |  |
| Others | 5 (6%) | 15 (17%) |  |
| Economic status |  |  | 0.251 |
| Low | 30 (34%) | 25 (29%) |  |
| Medium | 37 (43%) | 39 (45%) |  |
| High | 20 (23%) | 23 (26%) |  |
| Tobacco user |  |  | 0.104 |
| Yes | 60 (69%) | 55 (63%) |  |
| No | 27 (31%) | 22 (37%) |  |
| IL-1β (pg/mL) | 11.21 ± 8.46 | 5.99 ± 2.31 | **0.000** |
| TNF-α (pg/mL) | 63.37 ± 49.64 | 28.22 ± 9.96 | **0.000** |

BMI: Body mass index; IL-1β : Interleukin-1 beta; TNF-α : Tumour necrosis factor-alpha; SD: Standard deviation. Significant p values are shown in bold.

Serum levels of IL-1β and TNF-α were found significantly elevated in MDD patients compared with HCs (*p* < 0.05). Spearman’s correlation study demonstrated significant positive correlations between serum levels of IL-1β and Ham-D scores (*r* = 0.773; *p* = 0.000) and between serum levels of TNF-α and Ham-D scores (*r* = 0.400; *p* = 0.002) in patient group. Variations of serum IL-1β and TNF-α among the study population were shown by Fig. 1. Correlation between the analyzed parameter and the severity of depression was presented in Table 2. A scatter plot of serum IL-1β and TNF-α levels with Ham-D scores among MDD patients was shown in Fig. 2. From this illustration, we observed that females tended to show greater serum IL-1β and TNF-α levels compared with males when at higher Ham-D scores.

**Table 2.** Spearman’s correlation between analyzed parameter and severity score in depression.

| Correlation parameters | MDD patients (n = 87) |  |
| --- | --- | --- |
|  | r | p |
| IL-1β and Ham-D score | 0.773 | **0.000** |
| TNF-α and Ham-D score | 0.400 | **0.002** |

r: Correlation co-efficient; p: Significance. Significant p values are shown in bold.

**Fig. 1.**
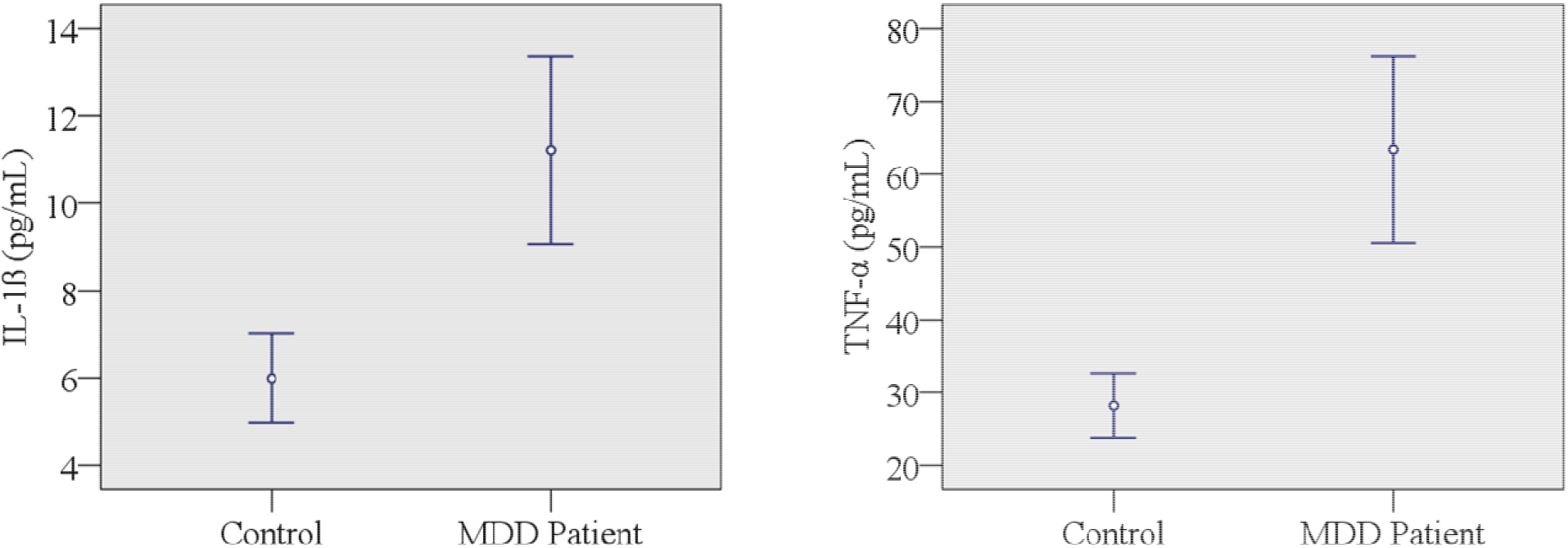
Variations of serum interleukin-1β (IL-1β) and tumor necrosis factor- α (TNF-α) among the study population. A significant difference between patient and control groups at a 95% confidence interval (CI).

**Fig. 2.**
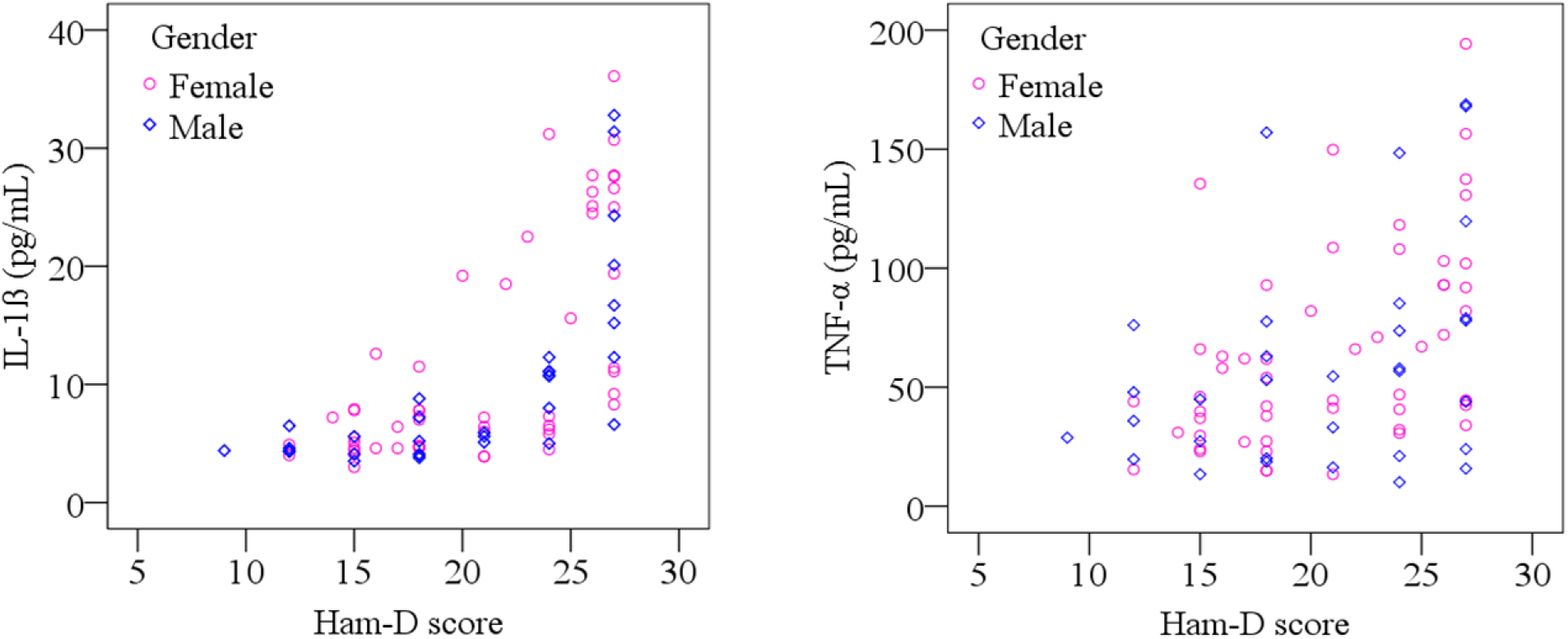
Scatter plot of serum interleukin-1β (IL-1β) and tumor necrosis factor- α (TNF-α) with Ham-D scores of MDD patients.

The cut-off points for the analytical purpose of serum IL-1β and TNF-α levels were detected as 6.05 pg/mL and 29.50 pg/mL, respectively by the ROC curve (Fig. 3 and Fig. 4). The area under the curve (AUC) for ROC analysis was 0.838 for IL-1β and 0.866 for TNF-α, both the results were significant at *p* values less than 0.001. Depressive conditions were indicated by the higher values. Sensitivity, specificity, positive predictive value (PPV) and negative predictive value (NPV) were 72.4%, 73.1%, 72.9% and 72.6% for IL-1β, whereas the values were 81.6%, 66.7%, 71.0% and 78.4% for TNF-α, respectively.

**Fig. 3.**
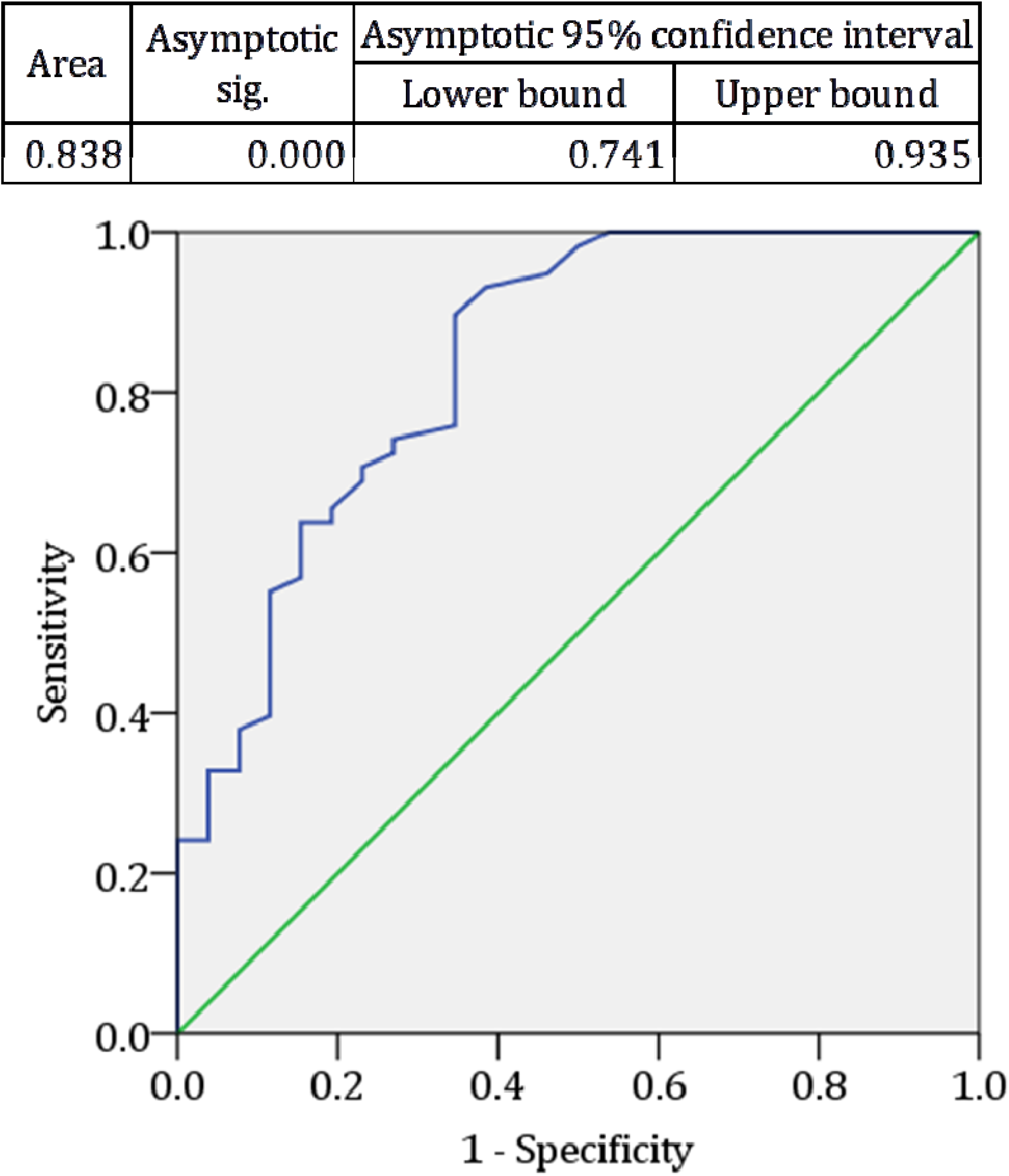
Receiver operating characteristic (ROC) curve for interleukin-1β (IL-1β. The cut-off point was detected as 6.05 pg/mL.

**Fig. 4.**
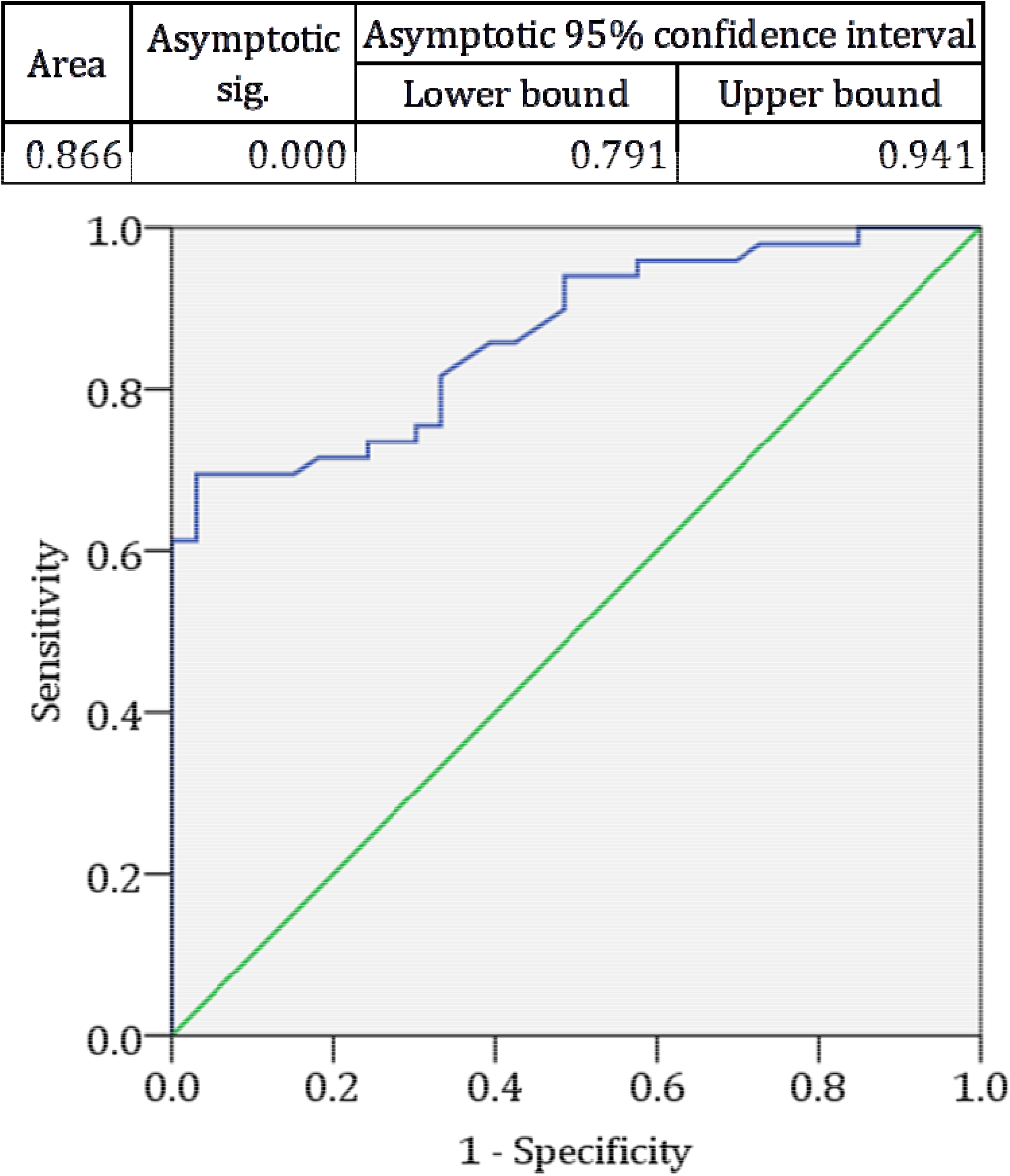
Receiver operating characteristic (ROC) curve for tumor necrosis factor- α (TNF-α). The cut-off point was detected as 29.50 pg/mL.

## 4. Discussion

The present study compared the serum levels of IL-1β and TNF-α among MDD patients and their corresponding HCs. Firstly, we identified the majority of MDD patients were females (63%), jobless (52%), and medium economic class (43%) which are consistent with several previous reports (Farid et al., 2007; Yang et al., 2007). According to the current findings, significant elevations of serum IL-1β and TNF-α levels were observed in MDD patients compared to HCs. However, some current literature indicating the inconsistencies with the present findings whether MDD agrees with the increased or no alterations of these cytokine levels. Few studies found serum IL-1β and TNF-α levels are not linked with MDD (Farid et al., 2007; Myung et al., 2016; Tsuboi et al., 2018; Yang et al., 2007).

Our results can be explained by different psycho-neuro-inflammatory mechanisms. Inflammatory cytokine IL-1β is a potent activator of the HPA axis, which is elevated in depressed patients (Berkenbosch et al., 1987; Sapolsky et al., 1987; Turnbull and Rivier, 1999). Increased serum levels of IL-1β has a major role in releasing hypothalamic corticotropin-releasing hormone (CRH), pituitary adrenocorticotropic hormone (ACTH), and adrenal steroidogenesis which cause depression via indirect mechanisms (Goshen and Yirmiya, 2009; Hsieh et al., 2010; Maes et al., 1993; Tominaga et al., 1991). TNF-α is produced by the actions of macrophages, natural killer cells, and T-lymphocytes (Han and Yu, 2014). HPA axis is then activated by the inflammatory response system (IRS) due to the production of TNF-α. The turnover of catecholamine and serotonin are increased as CRH and ACTH are produced previously. The cytokine, TNF-α reduces the uptake of serotonin from synaptic cleft thus produces depressive symptoms (Maes, 2011; Ng et al., 2018). The homeostasis of the neurotransmitter serotonin (5-HT) can be destroyed by the elevated level of TNF-α thus depressive disorder may occur (Nutt, 2008). TNF-α increases dopamine metabolism which results in depression symptoms (Van-Heesch et al., 2013). According to the past studies, elevated levels of cytokines are considered as major causes for developing depression and there is a link existing between psychological stress and MDD (Kim et al., 2016; Iwata et al., 2013).

The present study findings are supported by many previous research outputs. Studies reported that patients with MDD had significantly higher levels of IL-1β, IL-10, and TNF-α compared to HCs (Sadia et al., 2020; Zou et al., 2018). Another study by Leo et al. proposed that MDD patients had elevated baseline values of IL-1β, IL-6, and TNF-α compared with healthy subjects (Leo et al., 2006). Mota et al. observed the higher levels of serum IL-1β in major depressive patients than controls (Mota et al, 2013). Another study detected the higher plasma levels of IL-1β in MDD patients compared to HCs (Miklowitz et al., 2016). The baseline serum levels of IL-6 and IL-1β were significantly elevated in the MDD compared to the control group (Zhang et al., 2018). Tuglu et al. observed elevated serum TNF-α levels in MDD patients compared to control subjects (Tuglu et al., 2003). Along with TNF-α, serum IL-6 and IL-18 were found significantly increased in MDD (Al-Hakeim et al., 2015). Mikova et al. also reported higher serum TNF-α levels among depressed patients (Mikova et al., 2001). According to Fan et al. MDD patients had increased serum TNF-α than healthy volunteers (Fan et al., 2017).

We observed a significant positive correlation between analyzed cytokines and severity of depression which is consistent with several past studies (Madi et al., 2017; Tsuboi et al., 2018). Moreover, we observed females with higher Ham-D scores showed greater serum IL-1β and TNF-α levels compared to males. Likewise, ROC analysis showed significant predictive values of increased serum IL-1β (AUC: 0.838, CI: 0.741-0.935, and *p* < 0.001) and TNF-α (AUC: 0.866, CI: 0.791-0.941, and *p* < 0.001) levels. Additionally, we noticed MDD patients with high Ham-D scores (≥ 25) were females, jobless, and belong to the poor socio-economic class. However, we did not record any food intake data and duration of MDD symptoms of the study population; these can be considered as limitations of the present findings.

## 5. Conclusion

To the best of our knowledge, this is the first-ever study examining the association of serum IL-1β and TNF-α levels with major depression among the Bangladeshi population. The increased probability of developing MDD may be associated with elevated serum levels of IL-1β and TNF-α. We believe these results can be used as predictors for the assessment of depression risk. Further studies with a large and more homogeneous population are required to understand the exact biological mechanisms through which the measured cytokines are linked to depression.

## Data Availability

Data supporting this research will be available from the corresponding author upon reasonable request.

## Acknowledgments

All the authors are thankful to all the staff and physicians who worked at the department of psychiatry, BSMMU, Dhaka, Bangladesh for their technical and administrative support. The authors are also thankful to the Institute of Nutrition and Food Science, University of Dhaka, Dhaka, Bangladesh for providing laboratory support.

